# Quantitative Grey/White Matter Myelin differences in Neurofibromatosis Type-1 using T1W/T2W ratio

**DOI:** 10.1101/2025.03.28.25324840

**Authors:** Varun Arunachalam Chandran, Caroline Lea-Carnall, Stavros Stivaros, Grace Vassallo, Nils Muhlert, Shruti Garg

## Abstract

**Background:** Neurofibromatosis-1 (NF1) is an autosomal dominant neurodevelopmental condition commonly characterised by learning difficulties, with co-occurring autism spectrum conditions in 30% and attention deficit hyperactivity disorder in about 50% of affected school-age children. The structural brain phenotype characteristically shows T2-white matter hyperintensities, particularly in the thalamus and basal ganglia, and previous diffusion MRI studies have demonstrated widespread white matter microstructural differences, suggesting aberrant myelination in NF1. However, no previous studies have investigated quantitative myelin in NF1. The aim of this study is to compare quantitative myelination in NF1 compared to neurotypical controls, and examine the relationship between myelin and working memory performance in children with NF1.

**Methods and materials:** The T1-weighted and T2-weighted structural MRI images were acquired from 48 children and adolescents with NF1 aged 11-18 years. These were compared to data from 168 age- and gender-matched neurotypical controls drawn from the Human Connectome Project. Visuospatial n-back tasks were used to measure the working memory performance in the NF1 cohort. Pituitary depth was used as a covariate to control for pubertal differences.

**Results:** Compared to neurotypicals, children with NF1 showed significantly reduced grey matter/white matter myelin ratios both throughout the brain and within each of the four major cerebral lobes. The stratified age-related whole-brain myelin differences were reduced in NF1 relative to controls. The sex-related whole-brain myelin differences were found to be greater in females than in males. However, no significant correlation was found between myelin and working memory performance in NF1. Pituitary depth was reduced in NF1 relative to controls.

**Conclusion:** The reduced grey matter/white matter (GM/WM) myelin ratios in the whole brain and the frontal, temporal, parietal and occipital lobes suggests a widespread disruption of cerebral myelination in NF1. The age and sex-related myelin differences, and associated differences in developmental trajectories, may occur due to the effects of gonadal hormones but needs to be examined in future studies.

## Introduction

Neurofibromatosis-1 (NF1) is an autosomal dominant tumour predisposition syndrome which affects 1 in 3000 people (Lee et al., 2023). Cognitive and behavioral difficulties are present in 70% of children with NF1 (Hachon et al., 2011), while autism spectrum conditions co-occur in 10-30%, and ADHD in 50% of cases. NF1 is caused by a *de-novo* or inherited mutation of the NF1 gene, which primarily regulates the activity of the Ras/MAPK and downstream pathways. NF1 is highly expressed in oligodendrocytes which are myelin-producing glial cells (Asleh et al., 2020). Loss of NF1 has been shown to affect myelin integrity (karlsgodt et al., 2012), which may contribute to the learning impairments seen in NF1 (Garg et al., 2013). In this work, we use advanced neuroimaging methods to infer levels of myelin in the brain and then investigate whether this is associated with cognitive dysfunction.

Myelin plays a prominent role in learning and information processing speed in the brain. The myelination process begins during the second trimester of foetal brain development and persists into the second year of childhood (Jakovcevski et al., 2009). Myelination continues to develop and influence information processing speed rates from early childhood to young adulthood (Yeung et al., 2014). Cortical myelination (estimated using myelin water fraction and quantitative-T1 imaging) appears to increase with age (1-6 years) in the brain, including the inferior frontal gyrus, precentral gyrus and inferior temporal gyrus (Deoni et al., 2015). Similarly, in infants and toddlers, white matter myelination (estimated using myelin volume fraction) has been found to increase with age in the anterior corpus callosum, left inferior frontal gyrus and bilateral corona radiata (O’Muircheartaigh et al., 2014). These age-related changes in myelination have clear cognitive correlates, with increasing myelin water fraction in the temporal and frontal cortices during early childhood linked to improvements in expressive and receptive language. Myelin progression in the grey and white matter of the cerebrum during childhood and adolescence may therefore play a crucial role in acquiring various cognitive abilities (Corrigan et al., 2021).

Studies using diffusion tensor imaging show aberrant white matter microstructure linked to myelin differences in the major fibre tracts including anterior thalamic radiation, inferior fronto-occipital fasciculus and corpus callosum underpinning learning difficulties in NF1 (Karlsgodt et al., 2012, Ferraz-Filho et al., 2012). Executive function in NF1 has also been linked to reduced fractional anisotropy and increased mean diffusivity in the anterior thalamic radiation, cingulum bundle and superior longitudinal fasciculus (Koini et al, 2017). In particular, white matter abnormalities in the anterior thalamic radiation were strongly related to deficits in inhibitory control in NF1. Some previous studies have shown co-occurrence of grey matter volume and white matter microstructure differences affects myelin (Nemmi, et al., 2019, Karlsgodt et al., 2012), however the quantitative myelin in the grey matter and white matter and its impact on learning in NF1 remains unknown.

Neuroanatomical studies report focal areas of high signal intensity on T2-Weighted MRI images in 60-70% of children with NF1 (Han et al., 2024, Payne et al., 2014). These T2 hyperintensities (T2H) are predominantly seen in the subcortical structures including the thalamus, basal ganglia, brainstem, and cerebellum (Goh et al., 2004, Piscitelli, et al., 2012). They exert no mass effect and are not associated with any focal neurological deficit. They begin arising typically in early childhood and appear to resolve by late adolescence, thought to be due to repair or reorganisation of myelin (Baudou et al., 2021, Hyman et al., 2003). The relationship between T2H and cognition is unclear and the findings are contradictory. Whilst some studies have found no relationship between T2H and cognition (Moore et al., 1996, Bawden et al., 1996), others have shown a relationship between the number and location of T2H with cognition (Chabernaud et al., 2009, Feldmann et al., 2003).

The T2H, along with changes in the white matter tracts and the microstructural deficits described by diffusion MRI studies in NF1, are thought to be due to myelin abnormalities, including increased vacuolation, delay in myelination and demyelination (de Blank et al., 2023). But this evidence is predominantly derived from animal models of NF1. In humans, NF1 studies have reported brain structural differences, but not yet linked these changes to alterations in myelin. To address this key gap, we sought to examine the GM and WM myelin in NF1 compared to neurotypical controls. To achieve this, we used the T1W/T2W ratio derived from conventional T1- and T2-weighted structural brain images to obtain an indirect measure of quantitative myelin. We hypothesized that the T1W/T2W ratio in grey and white matter will be reduced in NF1 as compared to neurotypical controls. Further, we investigated the relation between cortical myelin and learning and predicted that reduced myelination in NF1 will be related to worse performance on executive function tasks.

## Methods and materials

### Participants

Forty-eight children and adolescents (age: 11-17 years old; gender: both males and females) with NF1 were recruited from the Manchester Centre for Genomic Medicine NF1 clinics and through NF charities newsletters and social media pages. Inclusion criteria were clinical diagnosis made using the National Institute of Health diagnostic criteria and/or molecular diagnosis of NF1 (Legius et al., 2021). The exclusion criteria included no history of brain injury/intracranial pathology other than asymptomatic optic pathway or other asymptomatic and untreated NF1-associated white matter lesion or glioma; (iii) no history of epilepsy or any major mental illness and (iv) no MRI contraindications. Written informed consent was provided by the parents of all children with NF1 recruited in this study. The study was initiated after receiving approval from the NHS research ethics committee.

### Working memory assessment

The verbal and visuospatial n-back task was used to assess working memory performance in all NF1 participants. The n-back working memory task was designed using the E-prime stimuli presentation software. These n-back tasks were applied at four different levels including 0-back, 1-back, 2-back, and 3-back with the task complexity increasing at each stage. The visuospatial n-back task used the blue squares presented on a black grid (3×3), except for the centre position. Participants were instructed to hit the key ‘A’, whenever they see the target that matches the corresponding squares in accordance with various levels of complexity. We used 0-back and 3-back accuracy and response times to compare low and high working memory load performance with myelination. Accuracy was measured as a proportion of correct hits and correct misses corresponding to each trial within a block (averaged across each n-back condition), while the response time was measured as the time lapse from the beginning of each trial and time taken to hit the button. Vineland Adaptive Behavioural Scale (VABS-III), a parent-rated questionnaire, was administered to assess the communication, daily living skills, socialisation and adaptive behaviour composite scores (Carter et al., 1998).

### MRI data collection

Whole brain T1- and T2-weighted structural brain images were acquired using the Philips 3T MRI scanner based in the Clinical Research Facility, Manchester. The structural MRI acquisition protocol included are as follows: T1-weighted structural MRI images: 3D-Fast field echo with TR= 8.40 ms, TE= 3.84 ms, voxel= 0.94 × 0.94 × 1 mm^3^, matrix= 256 × 256 mm^2^, FOV= 240 × 240 mm^2^, flip angle=8°, and slices=160. T2-weighted structural MRI images: Turbo Spin echo with TR= 3756 ms, TE= 88 ms, voxel= 0.45 × 0.45 × 4 mm^3^, matrix= 512 × 512 mm^2^, FOV= 230 × 183 mm^2^, flip angle= 90°, and slices= 40. A separate sample of one-hundred and sixty-eight neurotypical controls with whole brain T1-weighted and T2-weighted structural brain images (age: 132-215 months, gender: both males and females) from the Human Connectome Project (https://www.humanconnectome.org/study/hcp-lifespan-development), were used for comparison.

### T1W/T2W myelin mapping

T1W/T2W ratio is a quantitative myelin mapping technique used to measure the myelin concentration and tissue microstructure (Glasser & Van Essen, 2011, Uddin et al., 2019). T1W/T2W ratio provides an enhanced contrast to noise ratio in measuring both grey and white matter myelin. This myelin mapping technique is comparable to imaging methods like myelin water fraction and magnetization transfer imaging (Pareto et al., 2020, Ganzetti et al., 2014). Unlike other quantitative myelin imaging techniques, T1W/T2W myelin mapping approach takes the advantage of using conventional T1- and T2-weighted structural MRI brain images acquired for routine clinical practice and takes less time without having to apply advanced imaging protocols.

### Image preprocessing

The T1- and T2-weighted structural brain images from both NF1 and Human Connectome Project datasets were preprocessed using the FSL analysis suite (https://fsl.fmrib.ox.ac.uk/fsl/fslwiki/FSL). First, the T1W and T2W structural brain images were subjected to bias field correction. Then, the T2W images were coregistered to T1W images using normalised mutual information (Jenkinson, M., & Smith, S. 2001). The T1W/T2W was computed to generate whole-brain myelin maps. The segmented GM and WM brain images were applied to the T1W/T2W myelin maps to mask out the GM and WM specific T1W/T2W maps. Likewise, GM/WM values for the frontal, temporal, parietal lobe and occipital lobe were extracted using the MNI structural atlas.

Pituitary depth was used as a proxy measure for puberty (Sari et al., 2014). Pituitary depth was measured using the T1-Weighted structural MRI brain images from a sagittal plane, with clear visibility of the cerebral aqueduct, with the application of RadiAnt DICOM viewer. Subsequently, a scale was placed covering the superior and inferior end to measure the depth of the pituitary gland.

### Statistical analysis

Using R studio, Analysis of Covariance (ANCOVA) was applied to examine the whole brain GM/WM myelin and lobes derived from the T1W/T2W ratio between NF1 and control groups. Further, whole-brain myelin was tested separately between three different age-groups (11-13, 13-15, 15-18 years) in NF1 and neurotypical controls. Age, sex and pituitary depth were included as covariates. Similarly, whole-brain myelin was tested for sex differences in males and females separately between three different age groups between NF1 and neurotypical controls. Age and pituitary depth were included as covariates. Pearson’s partial correlation was applied to test the relationship between the GM/WM myelin and working memory performance in NF1 including age and gender as covariates. All statistical analyses were subjected to Bonferroni correction for multiple comparisons.

## Results

There were no significant age or sex differences between the NF1 and control groups. The pituitary depth was found to be significantly reduced in NF1 compared to neurotypical controls. Our results showed a highly significant reduction in GM/WM myelin ratios at the whole-brain level in NF1 compared to controls. Lobar-level analysis was then performed to examine myelin differences in frontal, temporal, parietal and occipital lobes. We found significantly reduced GM/WM myelin ratios in NF1 relative to controls across each of the four lobes (Table: 2, Fig. 2).

**Fig. 1:**
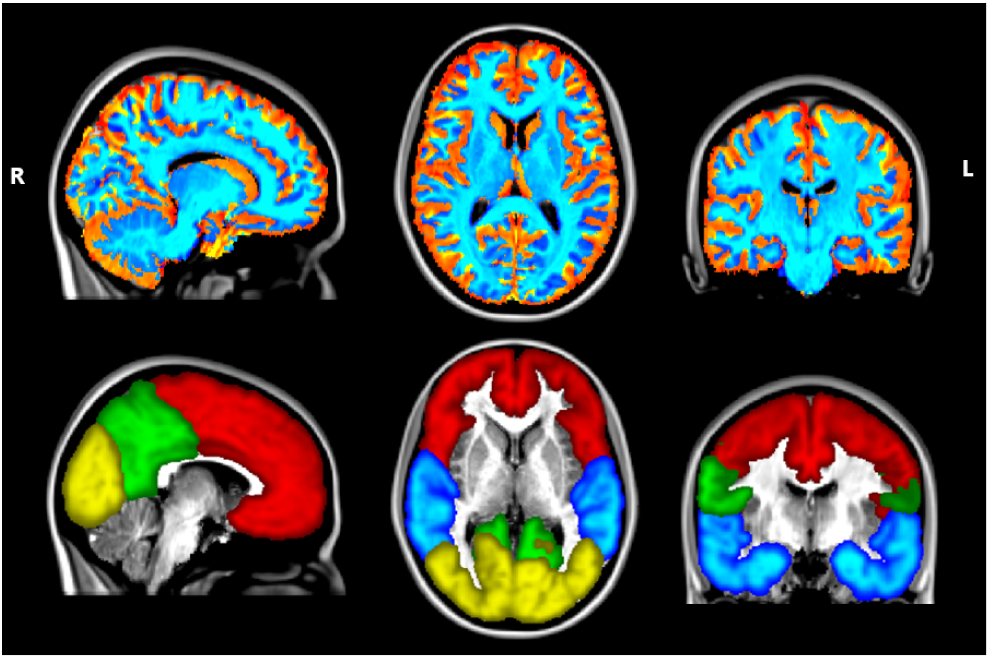
Representation of GM/WM myelin map in the whole brain and lobes. T1W/T2W grey matter (red-yellow) and white matter map (light-blue) from an individual with NF1 overlay on MNI standard brain template (top), the frontal (red), temporal (blue), parietal (green) and occipital lobe (yellow) masks overlay on the T1W/T2W map (bottom).

**Fig. 2:**
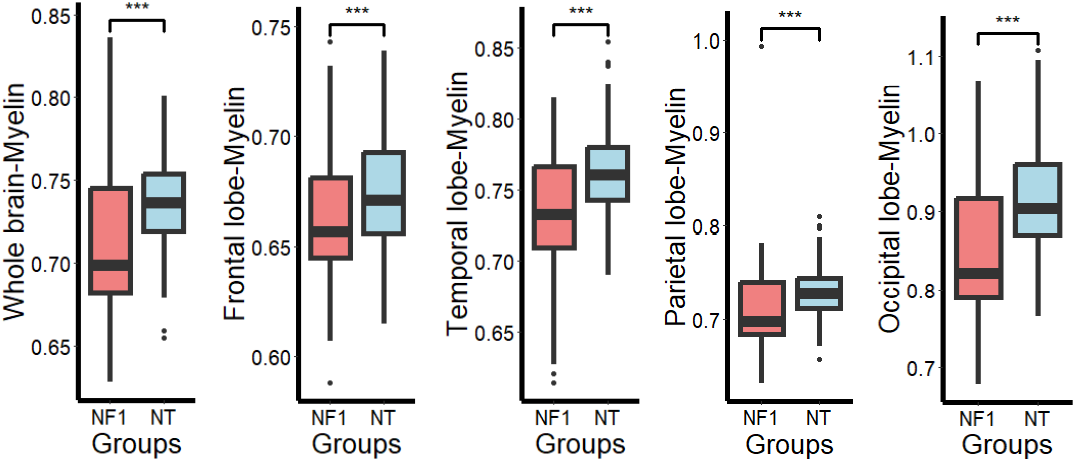
Comparison of GM/WM myelin differences between NF1 and neurotypical controls. Boxplots comparing whole-brain, frontal, temporal, parietal and occipital lobe myelin between NF1 and controls.

We then compared age-related changes in NF1 as compared to the control groups. For this purpose, we divided the age range from 11-13, 13-15 and 15-18 years. Whole-brain myelin differences were significantly different between the groups across all age bands (Table 3, Fig. 3). We also observed significant whole-brain myelin differences at 15-18 years in NF1 males compared to controls (Table: 4, Fig.4). In contrast, the females with NF1 showed significant whole-brain myelin differences at 11-13 years and 13-15 years (Table: 5, Fig.4).

**Table 1:** Participant characteristics. N-Number of participants, SD-Standard Deviation, M-Male; F-Female, N/A-Not Applicable, NT-Neurotypical Controls, VABS-Vineland Adaptive Behaviour Composite, WM-Working Memory, Acc-Accuracy

| Variables | NF1 (N=48) |  | NT (N=168) |  | P-value |
| --- | --- | --- | --- | --- | --- |
|  | Mean | SD | Mean | SD |  |
| Age | 14.67 | 1.894 | 14.15 | 1.687 | 0.070 |
| Gender (M/F) | 25/23 |  | 81/87 |  | 0.636 |
| Pituitary depth | 5.733 | 0.985 | 7.032 | 1.090 | <0.001 |
| VABS-ABC | 71.71 | 18.924 |  |  |  |
| WM-0 back (Acc) | 0.973 | 0.048 |  |  |  |
| WM-3 back (Acc) | 0.708 | 0.085 |  |  |  |

**Table 2:** Comparison of myelin between NF1 and neurotypical controls. GM-Grey Matter, WM-White Matter, F-effect size, ^*^*p* < 0.05, ^**^*p* < 0.01, ^***^*p* < 0.001

| Brain regions | NF1 (N=48) |  | NT (N=168) |  |  |  |
| --- | --- | --- | --- | --- | --- | --- |
| Whole brain level | Mean | SD | Mean | SD | F | P-value |
| GM/WM | 0.710 | 0.043 | 0.735 | 0.026 | 25.038 | <0.001*** |
| **Lobar level** |  |  |  |  |  |  |
| Frontal | 0.663 | 0.031 | 0.674 | 0.024 | 11.731 | <0.001*** |
| Temporal | 0.732 | 0.048 | 0.762 | 0.030 | 24.746 | <0.001*** |
| Parietal | 0.712 | 0.056 | 0.727 | 0.026 | 10.835 | 0.001*** |
| Occipital | 0.849 | 0.091 | 0.910 | 0.067 | 19.126 | <0.001*** |

**Table 3:** Comparison of whole brain myelin between NF1 and controls stratified across different age groups. GM-Grey Matter, WM-White Matter, F-effect size, \**p* < 0.05, \*\**p* < 0.01, \*\*\**p* < 0.001

| Brain regions | NF1 (N=48) |  |  | NT (N=168) |  |  |  |  |
| --- | --- | --- | --- | --- | --- | --- | --- | --- |
| Whole brain level | N | Mean | SD | N | Mean | SD | F | P-value |
| 11-13 years | 15 | 0.704 | 0.038 | 59 | 0.735 | 0.023 | 18.354 | <0.001*** |
| 13-15 years | 26 | 0.720 | 0.046 | 102 | 0.738 | 0.028 | 9.571 | 0.002** |
| 15-18 years | 23 | 0.711 | 0.048 | 69 | 0.739 | 0.027 | 5.346 | 0.039* |

**Table 4:** Comparison of myelin between NF1 and controls based on different age groups in males. GM-Grey Matter, WM-White Matter, F-effect size, ^*^*p* < 0.05, ^**^*p* < 0.01, ^***^*p* < 0.001

| Brain regions | NF1 (N=48) |  |  | NT (N=168) |  |  |  |  |
| --- | --- | --- | --- | --- | --- | --- | --- | --- |
| Whole brain level | N | Mean | SD | N | Mean | SD | F | P-value |
| 11-13 years | 9 | 0.727 | 0.029 | 24 | 0.739 | 0.024 | 1.811 | 0.189 |
| 13-15 years | 15 | 0.740 | 0.028 | 50 | 0.748 | 0.025 | 2.031 | 0.159 |
| 15-18 years | 10 | 0.726 | 0.054 | 40 | 0.745 | 0.024 | 1.828 | 0.047* |

**Table 5:** Comparison of myelin between NF1 and controls based on different age groups in females. GM-Grey Matter, WM-White Matter, F-effect size, ^*^*p* < 0.05, ^**^*p* < 0.01, ^***^*p* < 0.001

| Brain regions | NF1 (N=48) |  |  | NT (N=168) |  |  |  |  |
| --- | --- | --- | --- | --- | --- | --- | --- | --- |
| Whole brain level | N | Mean | SD | N | Mean | SD | F | P-value |
| 11-13 years | 6 | 0.671 | 0.024 | 35 | 0.733 | 0.023 | 32.302 | <0.001*** |
| 13-15 years | 11 | 0.693 | 0.053 | 52 | 0.728 | 0.029 | 9.343 | 0.003** |
| 15-18 years | 13 | 0.699 | 0.042 | 29 | 0.729 | 0.029 | 0.598 | 0.444 |

**Fig. 3:**
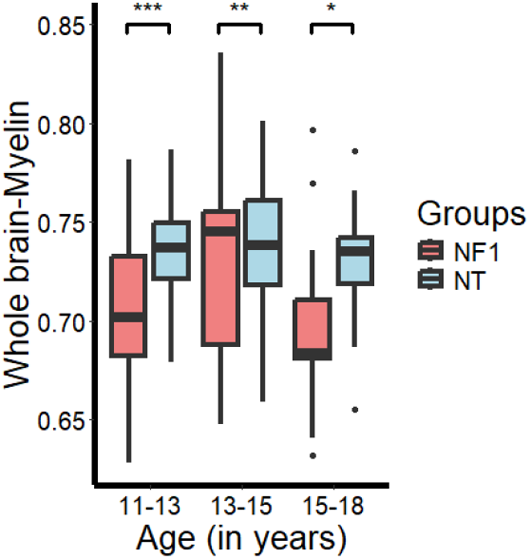
Comparison of whole brain myelin between NF1 and controls across different age groups. Boxplot showing stratified age-related whole brain myelin differences between NF1 and controls.

**Fig. 4:**
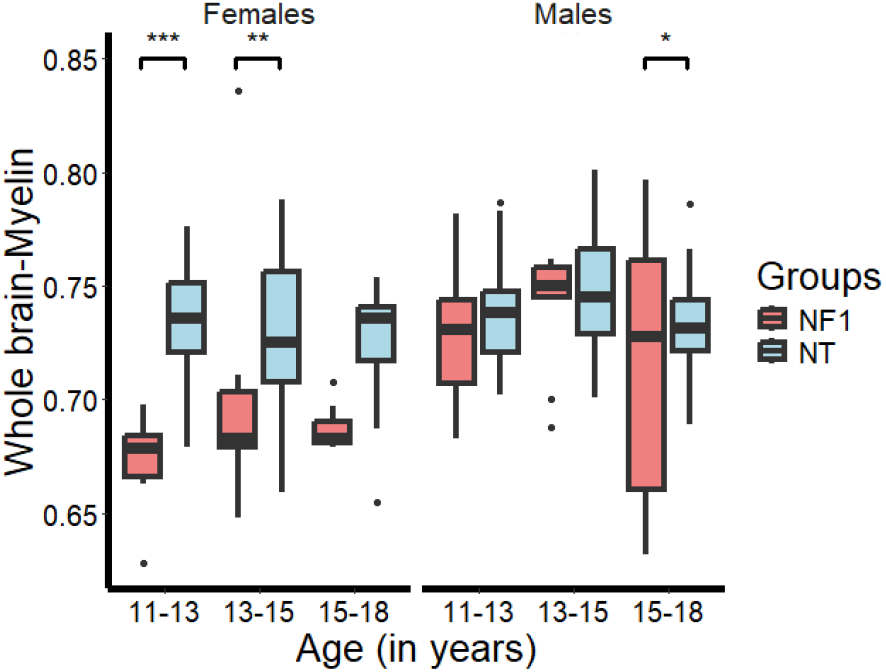
Relationship between myelin and different age groups in NF1 and controls (males and females) Boxplot graphical representation of sex-related (males and females) whole brain myelin differences between NF1 and controls.

Next, we investigated the relationship between myelin (whole brain and lobes) and working memory. NF1 participants completed 0-back and 3-back working memory tests, and accuracy and response times were analysed. We found no significant correlation between myelin and any of the working memory metrics (Table: 6).

**Table 6:** Correlation between myelin and working memory in NF1. ABC-Adaptive Behaviour Composite, Acc-Accuracy, RT-Reaction Time, r-Correlation

|  | Vineland-ABC |  | 0-back-Acc |  | 0-back-RT |  | 3-back-Acc |  | 3-back-RT |  |
| --- | --- | --- | --- | --- | --- | --- | --- | --- | --- | --- |
| Whole brain | r-value | p-value | r-value | p-value | r-value | p-value | r-value | p-value | r-value | p-value |
| GM/WM | 0.085 | 0.577 | -0.013 | 0.466 | 0.024 | 0.437 | 0.155 | 0.151 | 0.041 | 0.393 |
| **Lobar level** |  |  |  |  |  |  |  |  |  |  |
| Frontal | 0.023 | 0.883 | 0.080 | 0.298 | 0.167 | 0.134 | 0.124 | 0.207 | 0.208 | 0.083 |
| Temporal | -0.084 | 0.584 | -0.082 | 0.295 | 0.015 | 0.461 | 0.062 | 0.342 | 0.051 | 0.369 |
| Parietal | 0.098 | 0.520 | 0.027 | 0.430 | 0.080 | 0.300 | 0.238 | 0.055 | 0.134 | 0.188 |
| Occipital | 0.124 | 0.417 | -0.030 | 0.422 | -0.037 | 0.403 | 0.170 | 0.129 | -0.100 | 0.254 |

T2-white matter hyperintensities (WMH) were present in 58% (28/48) of all children and adolescent participants with NF1 as reported using the routine clinical structural brain MRI images. T2-WMH were commonly observed in the thalamus (8), basal ganglia (11), brainstem (7), corpus callosum (4) and cerebellum (13) in NF1 (Fig.5).

**Fig. 5:**
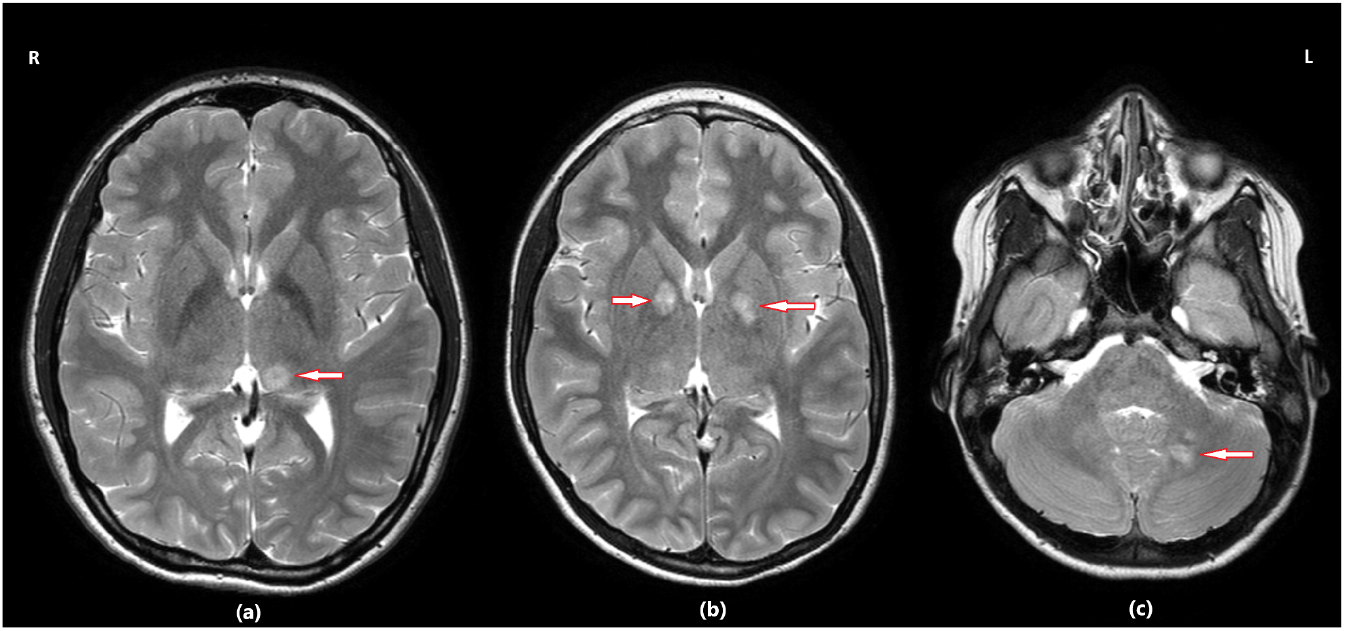
T2-white matter hyperintensity findings in NF1. The common areas of T2-white matter hyperintensities: (a) Left thalamus (b) bilateral globus pallidus (c) left cerebellar peduncle.

## Discussion

This is the first study, to our knowledge, to investigate quantitative MRI measures of myelin in individuals with NF1, utilizing the largest sample size to date. Our results demonstrated reduced myelin at a whole-brain level and in all four lobes in NF1 compared to controls. We also investigated age- and sex-related changes in myelination and found significant myelination differences in females with NF1 between the ages of 11-13 years and 13-15 years and in males between the ages of 15-18 years. However, quantitative myelin was not found to be associated with working memory performance or adaptive behaviour in NF1.

The lower myelin findings in the whole brain and cortical lobes in NF1 as compared to neurotypical controls are consistent with previous studies on *Nf1* animal models (López-Juárez et al., 2017, Mayes et al., 2013) and diffusion studies in NF1 clinical cohorts (Nemmi et al., 2019). Evidence from *Nf1* animal models suggests that myelin deficits in NF1 may be linked to aberrant neurobiology of oligodendrocytes and decompaction of myelin (Bennett et al., 2003). Neurofibromin protein, regulated by the NF1 gene, plays a key role in maintaining and regulating the RAS-MAPK signalling pathway, which in turn influences oligodendrocyte activity responsible for myelin production. Loss-of-function mutations of the NF1 gene and the resulting overactivation of RAS signalling pathway may contribute to the myelination differences seen in NF1 (Mayes et al., 2013). Similarly, diffusion studies in human clinical cohorts suggest myelin differences/axonal integrity in major white matter microstructure fibre tracts in NF1 (Baudou et al. 2020, Nemmi et al., 2019). T2 hyperintensities are commonly reported in NF1, most frequent in the thalamus and basal ganglia in children with NF1 (Greenwood et al., 2005). Previous studies based on post-mortem brain histological examination have suggested that myelin vacuolation/spongiform myelopathy may underlie T2H seen in NF1 (DiPaolo et al., 1995). Specifically, the fluid-filled vacuolar changes (intramyelinic edema) in the subcortical brain regions may be linked to NF1 myelin aberration. The GM/WM myelin deficits reported in our study support the role of aberrant myelin development and T2-white matter hyperintensities in NF1, and provide insight into the ages over which these changes become apparent.

The stratified age and sex-related myelin changes between children and adolescents with NF1 are consistent with white matter microstructure differences reported in previous studies (P De Blank et al., 2020, Tam et al., 2021). In a previous diffusion study imaging study, De blank et al (2020) found that children and adolescents with NF1 exhibited low fractional anisotropy, high mean diffusivity and radial diffusivity in the optic radiations, corpus callosum, frontal white matter tracts compared to controls, with these differences becoming more pronounced with age. Evidence from animal models suggest that aberrant glial cell proliferation/maturation may underlie these dynamic changes in myelin during young childhood and adolescence in NF1. T2 hyperintensities are highest in early childhood and may resolve over time, suggesting that the myelin differences represent atypical brain developmental trajectory in NF1 (Salman et al., 2018). Given that the young female adolescents and the old male adolescents with NF1 demonstrated lower myelin, this suggests that the sex differences may also play a significant role in GM/WM myelin development due to hormonal differences. Recent evidence from animal model studies indicates that the overactivation of Ras-MAPK signalling pathways-along with increased nitrous oxide levels may be driving the sex-specific differences in myelin abnormalities observed in NF1 (Hernandez et al. 2024). The inhibition of these hyperactive pathways led to the restoration of the myelin structure; but interestingly, these effects were not uniform across sexes. These results suggest that males and females with NF1 may respond differently to the dysregulation of these signalling pathways. Further, the sex hormones may interact with germline NF1 mutation through epigenetic mechanisms and produce changes in gene expression, affecting grey and white matter myelin development differentially (Diggs-Andrews et al., 2014).

A surprising finding in our study was that the pituitary depth was reduced in NF1 compared to neurotypical controls. In our analysis, the pituitary depth was used as a proxy measure to account for the effects of puberty. The pituitary gland plays an important role in puberty during adolescence. Specifically, the hypothalamus pituitary gonadal axis stimulates the production of sex hormones including testosterone and estradiol in males and females respectively beginning from early adolescence (Genc et al., 2023). These sex hormones are believed to accelerate the brain myelin maturation during puberty in adolescence (Herting et al., 2012). However, the sex hormones associated with pathophysiological differences in neurodevelopmental conditions like NF1 may affect the size of the pituitary gland during adolescence (Diggs-Andrews et al., 2014).

Contrary to our hypothesis, this study did not find a significant association between myelin and working memory performance in NF1. This may be partly explained due to our use of only a limited battery of cognitive tests. However, some previous studies using diffusion tensor imaging showed white matter microstructure differences linked to myelin deficits underpinning executive function difficulties in NF1(Nemmi et al., 2019, Karlsgodt et al., 2012). Further myelination differences have been noted in other neurodevelopmental populations including ASC and ADHD. Several studies using different structural neuroimaging techniques including magnetization transfer ratio (Gozzi et al., 2012), T1W/T2W ratio (Zhang et al., 2024), diffusion tensor imaging, myelin volume fraction (Lin et al., 2024) to assess the quantitative myelin and white matter microstructure in individuals with ASC and ADHD have shown diverse findings of myelin-related differences in frontal, temporal, parietal and occipital brain regions. The relationship between myelin and cognition in NF1 needs further investigation, particularly to examine the relationship between executive function and behavioural phenotypes of ASC and ADHD in NF1.

The application of T1W/T2W myelin mapping takes advantage of using high-resolution structural brain images without requiring the collection of data from advanced quantitative myelin imaging methods. The conventional structural MRI images were acquired within a very short time as compared to the advanced myelin imaging techniques, such that, the chances of having motion artefacts in the brain imaging data specifically from clinical and pediatric populations were minimised. This approach allowed us to use T1-weighted and T2-weighted structural brain images for neurotypical controls from large open-source cohorts. T1W/T2W myelin mapping is noted to be more sensitive to measuring GM than WM myelination (Sandrone et al., 2023). T1W/T2W mapping data must be interpreted with caution because the varying T1W and T2W signal intensities may sometimes mimic the biological properties of the tissue including iron, calcium and water concentration (Norbom et al., 2020). Validating the T1W/T2W study findings with other quantitative myelin techniques will improve reliability of this metric (Hagiwara et al., 2018, Arshad et al., 2017). Limitations of this study include relatively small sample size to investigate the stratified age- and sex-related myelin differences in NF1 compared to controls. These age and sex-related differences therefore need replication by larger studies using longitudinal designs to track changes over the developmental time course. Further we used T1-W and T2-W from open-source control images (from the Human Connectome Project) for case-control comparison. However, the controls were age and sex-matched, similar processing pipelines were used for analyses. This approach also allowed us to draw a control sample 3 times the size of the NF1 group.

## Conclusion

In summary, we found lower GM/WM myelin in the whole brain and the frontal, temporal, parietal and occipital lobes in NF1 adolescents compared to controls. Myelination differences remain consistent over age and indeed most prevalent in females and older males suggesting that sex may be a key factor influencing myelination. Our finding of reduced pituitary depth in NF1 compared to the neurotypical controls further reinforce the role of sex hormones in influencing myelination. Finally, our approach of using T1W/T2W ratio to identify quantitative myelin differences in NF1 may be applied to clinically acquired larger datasets.

## Data Availability

All data produced in the present study are available upon reasonable request to the authors

https://www.humanconnectome.org/study/hcp-lifespan-development

## Acknowledgements

VAC was funded by the NIHR Manchester Biomedical Research Centre (NIHR203308). The views expressed are those of the author(s) and not necessarily those of the NIHR or the Department of Health and Social Care”. SG was awarded a Francis Collins Scholarship to support this study through Neurofibromatosis Therapeutic Acceleration Program (NTAP).

## Authorship contributions

Varun Arunachalam Chandran: Formal Analysis, Investigation, Methodology, Writing-original manuscript.

Caroline Lea-Carnall: Writing – Methodology, Investigation and Writing – review & editing.

Stavros Stivaros: Investigation-Clinical evaluation

Grace Vassallo: Investigation

Nils Muhlert: Supervision, Methodology, Validation, Writing – review & editing.

Shruti Garg: Conceptualization, Supervision, Investigation, Writing – review & editing.

## Data sharing

Anonymized data used in this study can be made available upon request from the authors.

## Declaration

The authors declare that they have no known competing financial interests or personal relationships that could have appeared to influence the work reported in this paper.

